# Neural basis of negative future mental imagery-based simulation in the bipolar disorder spectrum

**DOI:** 10.1101/2024.10.29.24310750

**Authors:** Martina Di Simplicio, Renee M. Visser, Giorgio Alfarano, Julie L. Ji, Sana Suri, Emily A. Holmes

## Abstract

**Objectives:** Dysfunctional mental imagery-based simulations of emotional future events (episodic future simulation) - can contribute to emotional dysregulation in the bipolar disorder spectrum (BPDS). For example, vividly imaging social encounters having a disastrous outcome can trigger low mood. However, episodic future simulation may be perceptually modified to reduce their emotional intensity, which may in turn reduce BPDS mood instability. This study investigates the neural bases of perceptual and emotional characteristics of negative episodic future simulation in BPDS, and of perceptually manipulating such imagery.

**Methods:** Euthymic participants with BPDS (N=20) and healthy controls (HC, N=17) underwent an fMRI scan whilst generating, then perceptually modifying, negative episodic future imagery, and during rest.

**Results:** Negative episodic future imagery were rated as significantly more *real* and *unpleasant* in the BPDS compared to HC group. Critically, perceptual manipulation of such imagery reduced ratings in the BPDS group to HC levels. Whole-brain analyses of task-based neural activity revealed greater activation in networks including right frontal pole, middle and inferior frontal gyrus and insula during episodic future simulation in BPSD versus HC. BPDS participants showed greater intrinsic resting state functional connectivity within the default mode network than HC participants.

**Conclusions:** Findings suggest dysfunctional episodic future simulation circuitry across default mode network, fronto-parietal and fronto-insular areas may represent a neural mechanism via which mental imagery amplifies emotion in BPDS. Perceptual modifications to dampen such imagery successfully abolished subjective and neural differences between groups, indicating promise as an intervention tool targeting mood instability.

## 1. Introduction

Bipolar disorder is characterised by alternating periods of depression and (hypo)mania and an extremely high burden of disease, and is considered to be a severe form of mental disorder^1^. An estimated 2.4% of the global population live with a bipolar disorder spectrum (BPDS) diagnosis, which ranges from subclinical features of bipolar psychopathology, such as manic-like experiences and mood instability, to clear-cut (hypo)manic and depressive episodes^2^. Mood instability symptoms are very common and impair functioning even outside an illness episode^3^. Taking a spectrum approach of bipolar symptoms that includes such subclinical affect disturbance is thus key to improving our understanding of BPDS^4^. Psychological therapies for bipolar disorder remain unsatisfactory^5^, and mental imagery-based cognitive interventions have emerged as a promising approach for treatment development in this field^5^, in particular for mood instability^6,7^.

The role of dysfunctional prospection has been recently highlighted in psychopathology^8^. Prospection can include mental imagery-based representations of specific hypothetical future events, henceforth referred to as *episodic future simulation*, which has been proposed as a modifiable cognitive mechanism underlying mood instability in BPDS^9^. Mental imagery refers to internal perceptual experience in the absence of an external sensory input (“seeing in the mind’s eye”), manifesting as a weak form of sensory perception in the brain^10^. Mental imagery of future events, or episodic future simulation can allow us to “pre-experience” the event, eliciting emotional responses in an as-if-real manner, with downstream impacts on motivation and behaviour^11,12^. Evidence suggests that the perceptual characteristics of imagery contribute to the impact on emotions and behaviour^13^.

Individuals with BPSD report experiencing more intense episodic future imagery compared to healthy controls, including heightened perceptual characteristics (‘vividness’ or ‘realness’) and emotional responses^9,14^. Across mental disorders, greater vividness and realness of negative future imagery is associated with greater (sub)clinical mood instability^15,16^. Heightened perceptual strength of imagery could drive the emotional impact of mentally simulated negative future events and amplify symptoms in BPDS.

In the clinic, imagery is often felt by patients as reflecting external reality despite awareness of its being a form of internal cognition. Disrupting the perceived ‘realness’ of such imagery through modifying its perceptual characteristics or its content can reduce mood instability in BPDS^6^. Imagery-based cognitive therapy employs perceptual modification techniques to reduce the emotional impact of imagery via changing its sensory qualities (such as imagining the same event in black and white rather than in colour to make it less real)^7^.

Neuroimaging evidence suggests that BPDS psychopathology is underpinned by abnormal activity in the brain’s emotion regulation circuitry, including heightened limbic reactivity to emotional stimuli (e.g. external visual images)^17^. This suggests that people in the BPDS may have greater neural responses to emotional stimuli in general, including internal mental im-agery, but the neural basis of the perceptual strength and emotional impact of episodic future simulation in BPSD remains unknown. Moreover, dysfunctional resting-state brain connectivity has also been reported in BPSD^18^, including in the default mode network (DMN)^19^. As spontaneous episodic future simulation typically occurs when the mind is left wandering ‘at rest’^20^, it warrants exploring whether resting-state brain function alterations in BPSD may be related to spontaneous imagery, in particular whether imagery persists during rest after deliberately generating negative episodic future simulations.

In summary, identifying the neural basis of mental imagery-based episodic future simulation in BPDS is important to improve our mechanistic understanding of whether the subjective emotional impacts of such imagery is underpinned by aberrant emotional reactivity, by dysfunctional simulation circuitry, or both. Further, we use an experimental design during fMRI to test the impact of perceptual modification of mental imagery. Understanding whether perceptual modification of episodic future simulation can modify its emotional impact at the subjective and neural level will also help inform the use of perceptual modification techniques in treatment.

Thus, the present study aimed to investigate whether individuals with BPSD differ from healthy controls (HCs) in:

1. subjective experience and neural bases of episodic future simulation of negative events
2. subjective experience and neural bases of perceptual modification of episodic future simulation
3. resting state networks (RSN) functional connectivity, following episodic future simulation, in particular networks involved in spontaneous future simulation during mind-wandering

Specifically, we aimed to test the hypotheses that, relative to HCs:

1. individuals with BPDS would report greater intensity of subjective experience of episodic future simulation and associated abnormal neural activity
2. perceptual modification of episodic future simulation, but not simple repetition of such imagery, in BPSD would restore a) subjective perceptual characteristics and b) emotional impact of such imagery to HC levels.

As no previous neurofunctional investigation of episodic future simulation has been conducted in BPDS, we did not have specific localisation and directional hypotheses for brain activity and hence performed exploratory whole-brain analyses. However, based on episodic simulation research and the emotion regulation literature in BPDS, we expected that during episodic future simulation individuals with BPDS would likely present aberrant brain activity in areas that signal emotional reactivity^17^ and the valence of simulation, including limbic regions and the vmPFC, and/or in areas driving the vividness of future simulation, such as the hippocampus and/or occipito-parietal-cingulate cortex^21^. Hence secondary ROI analyses of these regions were also performed. Further, after negative episodic future simulation, individuals with BPSD may present greater RSN functional connectivity in the DMN (known to support spontaneously occurring future simulation) relative to HCs.

## 2. Methods and materials

### 2.1 Participants and procedure

Participants were recruited via advertisements in the community, public mailing lists, the Medical Research Council Cognition and Brain Science Unit (MRC CBSU) volunteers’ panel, and the Cambridgeshire and Peterborough NHS Foundation Trust. The study was approved by the NRES Committee East of England (Ref: 15/EE/019) and all participants provided their written and informed consent and were reimbursed for travel and time. The BPDS group included euthymic individuals on a spectrum of severity, defined as: (i) high levels of hypomanic experiences on the Mood Disorder Questionnaire (MDQ^22^) (MDQ scores ≥ 7) *and* a past major depressive episode on the Structured Clinical Interview for DSM-IV (SCID^23^), or (ii) bipolar disorder type 1, 2 or NOS on the SCID. Clinical screenings were administered by a trained psychiatrist. The control group included individuals without any lifetime psychiatric history (based on the SCID^23^) and with low levels of hypomanic experiences (MDQ scores < 3). Individuals with a family history of bipolar disorder were also excluded from the control group to control for the presence of a familial bipolar phenotype. Pre-screening was conducted via email or over the phone using an MRI eligibility questionnaire, the MDQ, the Quick Inventory of Depressive Symptomatology, Self-Report (QUIDS-SR^24^) and the Altman Self-Rating Scale for Mania (ASRM^25^) to assess current mood. Fifty-five right-handed participants eligible for MRI scanning, and with scores on the MDQ <3 or >7 were invited for screening. Our final sample with good quality fMRI data consisted of 20 participants in the BPDS group and 17 in the HC group. See Supplementary Material for full description of exclusion criteria.

### 2.2 Measures and task

#### 2.2.1. Questionnaires

##### Mental imagery

The following questionnaires were administered to assess mental imagery-based cognition, including spontaneous use of mental imagery (*Spontaneous Use of Imagery Scale,* SUIS^26^), *Spontaneous Use of Emotional Mental Imagery Scale,* E-SUIS^27^), deliberate mental imagery generation ability (*Vividness of Visual Imagery Questionnaire,* VVIQ^28^); *Plymouth Sensory Imagery Questionnaire,* Psi-Q^44^); and intrusive future imagery (*Impact of Future Events Scale,* IFES^29^). See details in Supplementary Materials.

##### Affect

The following questionnaires were administered prior to the fMRI scan to assess participants’ affective state: ASRM^25^, QIDS-SR^24^, *Beck Anxiety Inventory* (BAI^30^), *Multidimensional Assessment of Thymic States* (MAThyS^31^). See details in Supplementary Materials. In addition, visual analogue scale (VAS) ratings of sadness, irritability, anxiety, depression, elation and fearfulness were administered before and after the experiment. After the experiment participants also reported whether they had experienced any spontaneous imagery of the cue scenarios from the task (see below) during the resting state scans.

#### 2.2.2. Episodic Future Simulation Task

The Episodic Future Simulation Task (EFST) was performed inside the MRI scanner and involved participants generating mental imagery-based simulations of 20 negative future scenarios in response to standardised visually presented written scripts, see Figure 1. The scenarios depicted everyday situations such as being late for a meeting or attending an unfamiliar social situation, e.g.: “You arrive at the party alone/ There is music and people chatting/ You start walking around/ Realising you can’t see anyone you know, you feel sweat pouring down your back”. See Supplementary Materials for more details.

**Figure 1.**
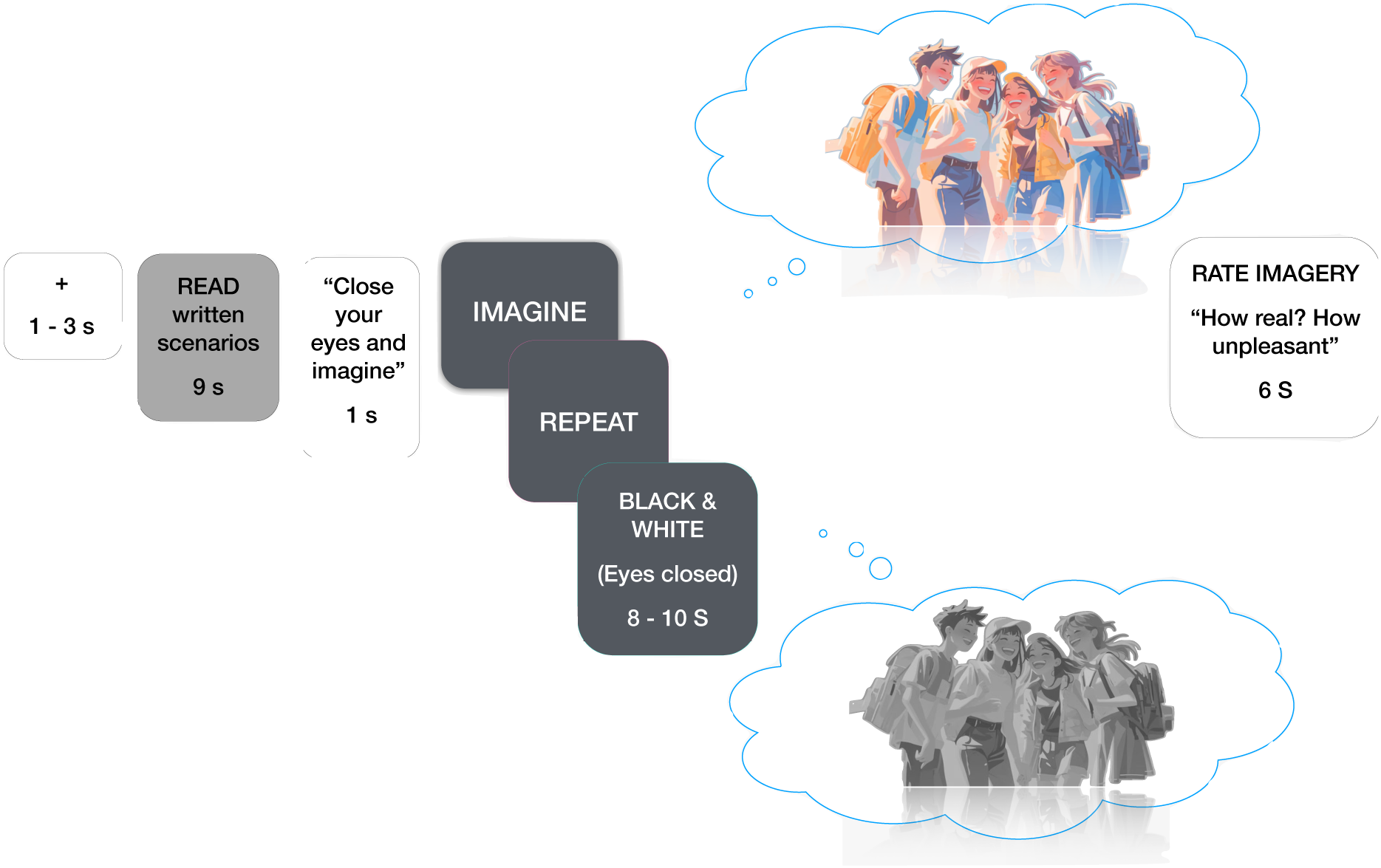
Episodic Future Simulation Task trial design. The figure depicts the structure of each single trial in the Episodic Future Simulation Task. Each trial presented: a fixation cross for 1-3 sec, followed by screen-displayed written descriptions of a negative scenario on a grey background screen for 9 sec, an instruction slide “close your eyes and imagine” (1 sec), and a blank slide for 8-10 sec during which participants imagined the scenario with their eyes closed. This was followed by an acoustic signal indicating the end of the trial and to open their eyes and instructions to rate the realness and unpleasantness of the imagined scenarios on a Likert scale from 0 to 10, using a button box. The instructions ‘Imagine’, ‘Repeat’ or ‘Black and White’ appeared at the start of each condition block, instructing participants to imagine (episodic future simulation condition), turn a previously imagined scenario black and white (perceptual manipulation condition) or repeat a previously imagined scenario (control condition).

##### 2.2.2.1. Episodic Future Simulation Task experimental conditions

The task had three within-subject experimental conditions: “Imagine”, “Black and White” (perceptual modification condition), and “Repeat” (control condition). All participants completed all three conditions. See Figure 2 for the experimental design.

**Figure 2.**
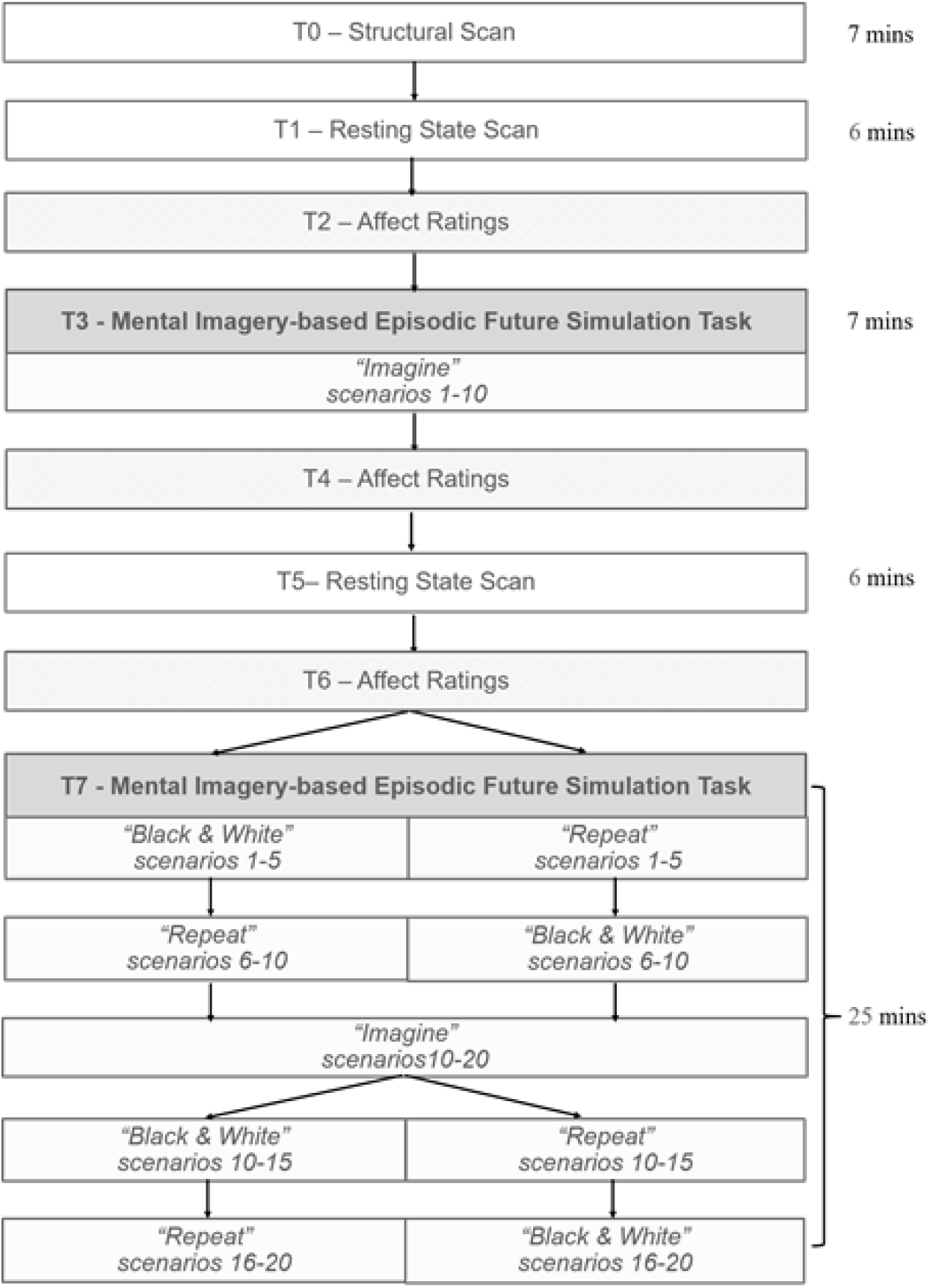
MRI session task sequence. The figure depicts every step in the MRI scan procedure with approximate duration.

###### Episodic Future Simulation

In the “Imagine” condition, participants were instructed to imagine the scenario for the first time as vividly as possible.

###### Perceptual Modification

In the “Black and White” condition, participants were instructed to bring to mind the previously imagined scenario while performing a perceptual modification involving turning the imagery from colour into black and white (like a sketch or a black and white movie; see Figure 1).

###### Control condition

In the “Repeat” condition, participants were instructed to simply imagine the previously imagined scenario again in exactly the same way. See Figure 1.

### 2.3 FMRI procedure and data acquisition

On arrival, participants completed the mental imagery and affect questionnaires and then practiced the EFST outside the scanner. After the practice, participants underwent a 3T brain MRI scan lasting approximately 45 minutes. The scan sequence included acquisition of a T1 (MPRAGE) structural scan (voxel=1x1x1mm), the EFST, and two close-eyed resting-state periods (EPI, voxel=3x3x3mm) (see Figure 2). At different points during the scan participants rated their happiness, sadness, and anxiety using three 11-point Likert scales (0= not all, 10=extremely). For data acquisition details see Supplementary Materials.

### 2.4 Analysis

The data that support the findings of this study are available from the corresponding author upon reasonable request.

#### 2.4.1. Questionnaires

Independent-samples t tests were used to compare questionnaire scores between the BPDS and the healthy control group. Mixed ANOVAs were used to compare VAS affect ratings before versus after the experiment (outside the scanner), and to compare Likert affect ratings at three time points during the scan (before simulation, T2, after simulation, T4, after rest, T6, see Figure 2), across the BPDS and HC groups. Significant interaction effects were decomposed by post-hoc within-group comparisons across time points.

#### 2.4.2. Behavioural data: Episodic Future Simulation Task

As realness and unpleasantness ratings were not normally distributed, non-parametric paired-samples Wilcoxon Signed Rank tests were used to compare these ratings between task conditions (Imagine vs. Repeat, Imagine vs. Black & White) and non-parametric independent samples Mann U Whitney tests were used to compare ratings between groups (BPDS vs. HC). Effect sizes were calculated using Rosenthal’s *r*.

#### 2.4.3. FMRI data

FMRI data processing was carried out using FSL v6 (FMRIB Software Library, Oxford Centre for Functional Magnetic Resonance Imaging of the Brain, Oxford University, Oxford, United Kingdom, http://www.fmrib.ox.ac.uk/fsl^32^).

##### 2.4.3.1. FMRI data: Episodic Future Simulation Task

See Supplementary Materials for description of data pre-processing.

###### First level analysis

In the first-level analysis, individual activation maps were computed using the general linear model with local autocorrelation correction. Five explanatory variables were modelled, including the different task components (read scenario, imagine, rate scenario, repeat imagery, turn imagery black & white, see Figure 1). The main contrasts of interest were Imagine vs. implicit baseline, Repeat vs. Imagine, and Black & White vs. Imagine. All variables were modelled by convolving the onset and duration of each stimulus with a haemodynamic response function, using a variant of a gamma function (i.e. a normalisation of the probability density function of the gamma function) with S.D.= 3 s and a mean lag of 6 s.

###### Group analysis

In the second-level analysis, whole-brain individual data were combined at the group level (BPSD vs HC) using a mixed-effects group cluster analysis corrected for multiple comparisons^33^. Significant whole-brain activations were identified using a cluster-based threshold of statistical images (height threshold of z= 2.3 and a (cluster corrected) spatial extent threshold of *p* < 0.05).

*A priori* analysis of regions of interest (ROIs) based on previous literature on episodic future simulation^21^ was conducted by extracting the BOLD signal change from anatomical masks of left and right hippocampus, medial prefrontal cortex (mPFC), and left and right amygdala^17^. Individual signal change was analysed using mixed-ANOVAs with Group as a between-groups factor and Task Condition as within-groups factor for the contrast of interest (Imagine versus baseline, Repeat vs Imagine and Black and White vs Imagine), with Bonferroni correction for three multiple comparisons. Accordingly, statistical significance was accepted at *p* < 0.017.

Associations between behaviour and neural activation were explored. Parametric modulation of neural activation by subjective ratings was tested by entering demeaned values for average realness and unpleasantness ratings per participant across groups during the Imagine condition (Imagine vs. implicit baseline). Individual signal change was extracted from areas of significant parametric modulation by ratings, and groups differences were explored using independent samples t-tests.

##### 2.4.3.2. FMRI data: resting state

See Supplementary Materials for description of data pre-processing.

Independent components analysis (ICA) was conducted using the FSL-MELODIC tool^34^. To concatenate components across all subjects, a group-level analysis across all participants was run identifying group-average components that can then be modelled for each individual subject. Because images were acquired from two repeated resting state scans (see Figure 2), both scans for each individual were used for the group ICA. A total of 25 components were identified in the group ICA and used to create subject-specific spatial maps and associated time series using dual regression.

Based on the literature on episodic future simulation, only the default network, frontoparietal network and sensori-motor network were analysed. To examine between-group differences in resting-state activity following future simulation, nonparametric permutation inference was employed using the FSL tool randomise, with 5000 permutations. Contrasts examined the main effects of time (before vs. after future simulation) and group (BPSD vs HC), as well as the interaction between time and group. Grey matter maps, generated via the FSL feat_gm_prepare script, were entered as voxelwise covariates of no interest. The resulting statistical maps were thresholded using Threshold-Free Cluster Enhancement. Where significant effects were found, the resulting clusters were thresholded correcting for multiple comparisons and used to generate masks to extract the mean time series from each participant to visualise results.

## 3. Results

### 3.1 Demographic and clinical characteristics

There were no significant differences between BPDS and HC groups on age, sex, level of education, marital status, occupational status or ethnicity. Demographic variables are reported in Table 1. Clinical characteristics of the BPDS group are reported in Supplementary Materials (Table 1S).

**Table 1.** Demographic characteristics of BPSD and HC groups.

|  | **BPSD (n=20)** | **HC (n=17)** | **$\chi^2 / t$** |
| --- | --- | --- | --- |
| **Sex, male (%)** | 9 (53) | 5 (25) | $\chi^2=.33$ p=.26 |
| **Age, years, M (SD)** | 29.6 (6.4) | 27.7 (3.9) | t=1.10, p=.28 |
| **Educational level, years, M (SD)** | 17.4 (2.7) | 17.4 (3.1) | t=.04, p=.97 |
| **Marital Status, single (%)** | 15 (75) | 10 (58.8) | $\chi^2=.29$, p=.24 |
| **Occupational status** |  |  |  |
| Employed full time | 5 | 10 | $\chi^2=.15$ |
| Employed part time | 2 | 3 |  |
| Unemployed | 2 | 0 |  |
| Student | 5 | 10 |  |
| **Ethnicity (% Asian vs White)** | 3 (17.7) | 3 (15) | $\chi^2=.76$, p=.55 |

### 3.2 Questionnaires

All results of between-group comparisons on baseline mental imagery and affect questionnaires and ratings are reported in Table 2.

**Table 2.**
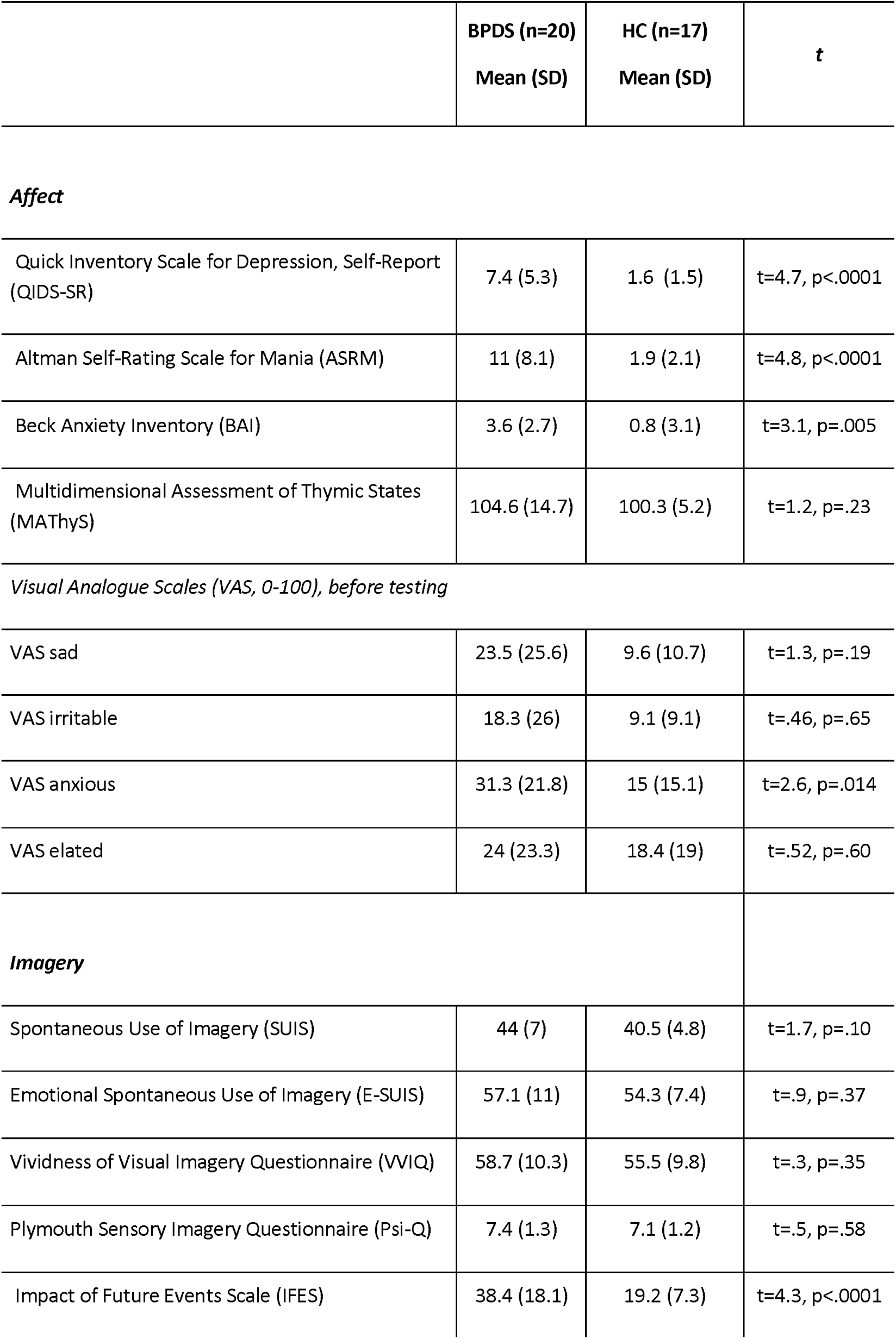
Questionnaires.

|  | **BPDS (n=20)**<br>**Mean (SD)** | **HC (n=17)**<br>**Mean (SD)** | **t** |
| --- | --- | --- | --- |
| ***Affect*** |  |  |  |
| Quick Inventory Scale for Depression, Self-Report (QIDS-SR) | 7.4 (5.3) | 1.6 (1.5) | t=4.7, p<.0001 |
| Altman Self-Rating Scale for Mania (ASRM) | 11 (8.1) | 1.9 (2.1) | t=4.8, p<.0001 |
| Beck Anxiety Inventory (BAI) | 3.6 (2.7) | 0.8 (3.1) | t=3.1, p=.005 |
| Multidimensional Assessment of Thymic States (MATHyS) | 104.6 (14.7) | 100.3 (5.2) | t=1.2, p=.23 |
| *Visual Analogue Scales (VAS, 0-100), before testing* |  |  |  |
| VAS sad | 23.5 (25.6) | 9.6 (10.7) | t=1.3, p=.19 |
| VAS irritable | 18.3 (26) | 9.1 (9.1) | t=.46, p=.65 |
| VAS anxious | 31.3 (21.8) | 15 (15.1) | t=2.6, p=.014 |
| VAS elated | 24 (23.3) | 18.4 (19) | t=.52, p=.60 |
| ***Imagery*** |  |  |  |
| Spontaneous Use of Imagery (SUIS) | 44 (7) | 40.5 (4.8) | t=1.7, p=.10 |
| Emotional Spontaneous Use of Imagery (E-SUIS) | 57.1 (11) | 54.3 (7.4) | t=.9, p=.37 |
| Vividness of Visual Imagery Questionnaire (VVIQ) | 58.7 (10.3) | 55.5 (9.8) | t=.3, p=.35 |
| Plymouth Sensory Imagery Questionnaire (Psi-Q) | 7.4 (1.3) | 7.1 (1.2) | t=.5, p=.58 |
| Impact of Future Events Scale (IFES) | 38.4 (18.1) | 19.2 (7.3) | t=4.3, p<.0001 |

Mixed ANOVAs comparing affect ratings on Likert scales at different time points during the neuroimaging scan (see Figure 2) showed a significant main effect of Group and a significant main effect of Time for anxiety ratings (group: *F* (1) = 13.60, *p* =.001; time: *F* (2.26, 76.8) = 5.12, *p* =.006). All participants were less anxious after the second resting-state scan (T3) compared to before task (T1), and individuals in the BPDS group were more anxious across the whole MRI scan. There was also a significant main effect of Time for happiness ratings (F (3,102) = 8.91, *p* < .0001) with all individuals being less happy at the end of the scan (T4) compared to before the task (T1). Finally, a significant two-way interaction between Group x Time was found for sadness ratings (*F* (2.38,81) = 3.40, *p* = .030): driven by participants in the BPDS group experiencing a marginal increase in sadness after the first “Imagine” task block (T2), followed by a significant reduction in sadness after the second resting-state scan (T4) (*F* (3) = 6.44, *p* = .001). No significant change in sadness was present in the control group during the whole scan period (*F* (3) =.54, *p* = .65).

### 3.3 Behavioural data: Episodic Future Simulation and Perceptual Manipulation

Participants in the BPDS group rated their episodic future imagery as significantly more real and more unpleasant compared to the HC group, both during the initial (Imagine; Real: Z=2.71, *p*=.007, *r*=.45; Unpleasant: Z=2.61, *p*=.009, *r*=.44) and repeated simulation (Repeat; Real: Z=2.34, *p*=.018, *r*=.39; Unpleasant: Z=1.90, *p*=.057, *r*=.32). Critically and as predicted, groups differences in subjective realness and unpleasantness were abolished when turning the simulations to black and white (Real: Z=1.15, *p*=.25, *r*=.19; Unpleasant: Z=1.68, *p*=.09, *r*=.28).

Critically, as predicted, across both groups, participants reported their episodic future simulation as less real (W=51.5, *p*<.0001, *r*=.73) and less emotionally unpleasant (W=33, p<.0001, *r*=.77) in the Black and White condition compared to the initial Imagine condition.

There was no significant difference in realness (W=298.5, p=.58, *r*=.09) and unpleasantness (W=337.5, *p*=.94, *r*=.01) ratings between the Repeat control condition compared to the initial Imagine condition. See Table 2S.

### 3.4 FMRI data

#### Episodic Future Simulation Task

##### Whole-brain analysis

Whole-brain analysis of task fMRI data revealed a significant cluster of deactivation for the interaction between group (BPDS vs HC) and task condition (Repeat vs Imagine). The cluster included voxels in the frontal pole (BA 9), inferior frontal gyrus (IFG, BA 8 and BA 46), middle frontal gyrus (MFG), orbitofrontal cortex and insula (Voxels=1572, Z=3.47, p=0.001, MNI peak= 20, 30, 36). This association was driven by individuals with BPSD showing greater activity (less deactivation) during the initial Imagine condition compared to the Repeat condition, while the opposite pattern was present in the HC group, see Figure 3.

**Figure 3.**
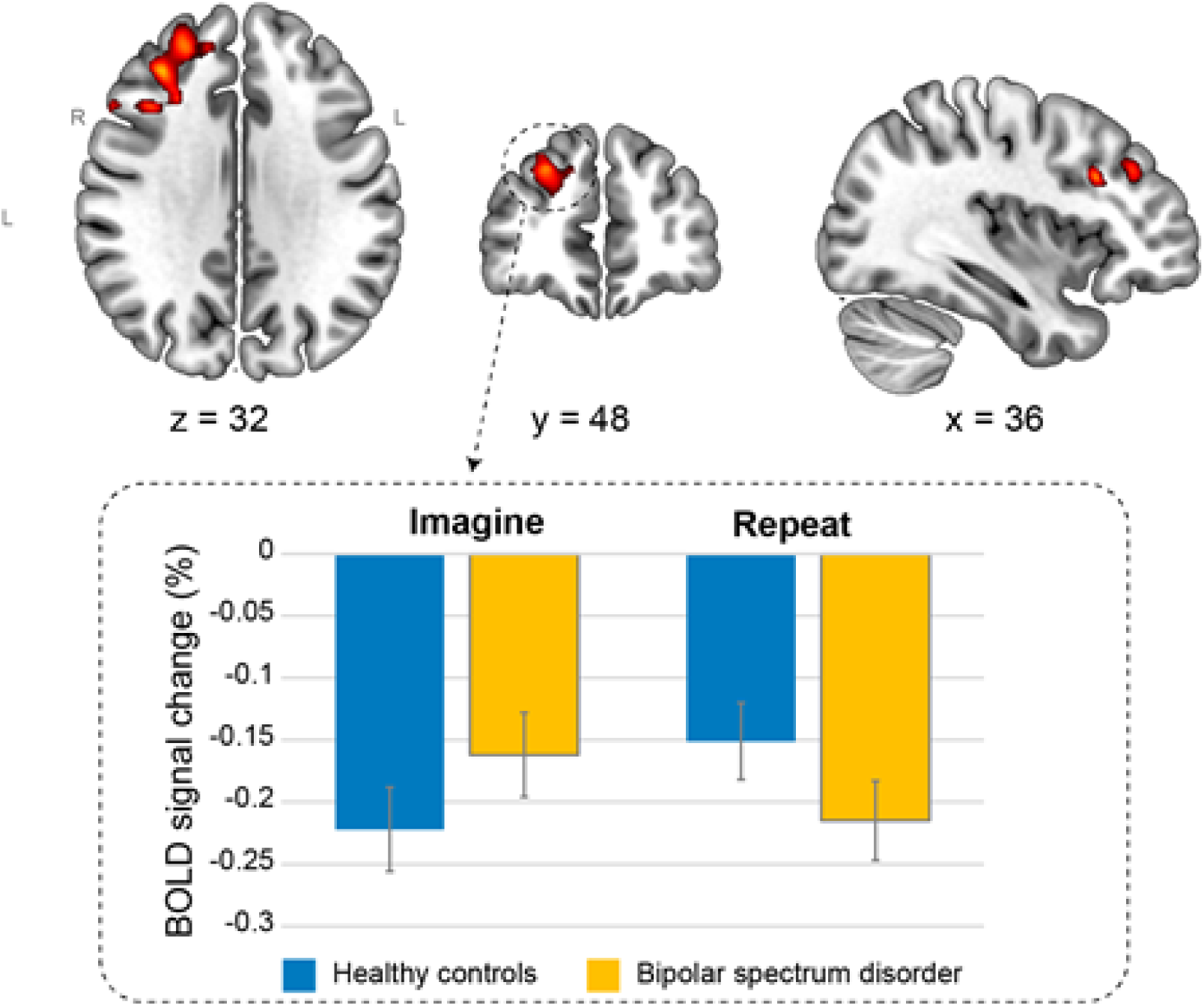
Heightened neural activity in BPSD vs HC during episodic future simulation. A whole-brain comparison between bipolar spectrum disorder (BPSD) and healthy control (HC) during repeated simulation of negative scenarios (Repeat) vs the first simulation of those scenarios (Imagine) reveals a cluster in the right middle frontal gyrus (1522 voxels; center of gravity: x=30.8, y=31.6, z=26.2; Z-max=3.47). Activation is thresholded using clusters determined by Z>2.3 (P<0.01) and a (corrected) cluster significance threshold of P=0.05. To illustrate where the differences originate from, the graph shows mean activation extracted from the whole cluster, per group, per task condition. L = Left, R = Right.

**Figure 4.**
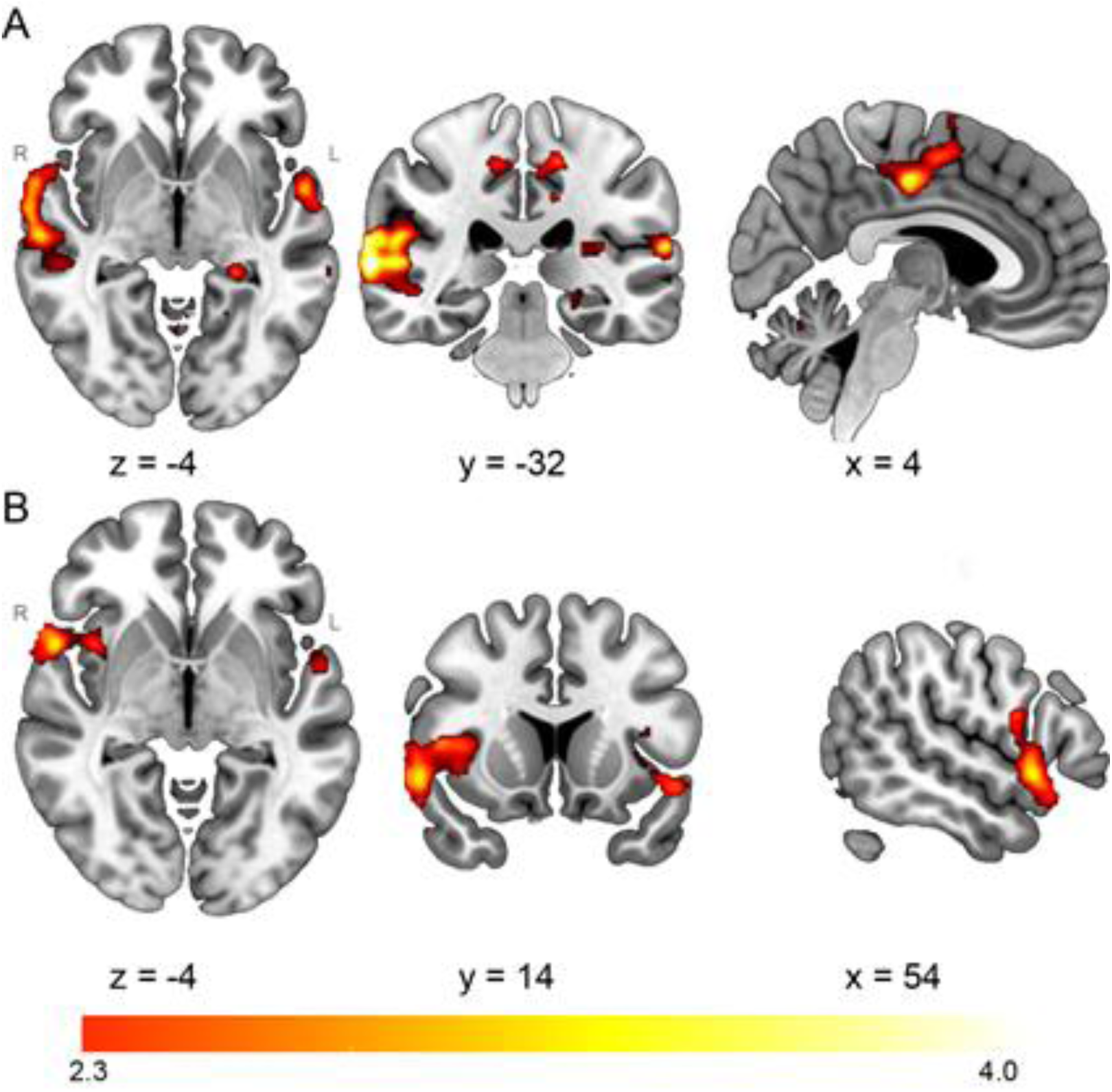
Neural activity of the core simulation network during episodic future imagery across the whole sample.

No other between group differences were present in any other contrast (Imagine vs baseline, Black and White vs Imagine).

##### ROIs analysis

There were no significant group x task condition interactions in any of the planned contrasts of interest in any of the a priori ROIs (medial PFC, bilateral amygdala and bilateral hippocampus).

##### Brain-behaviour correlations

Six clusters showed significant parametric activation associated with realness ratings during the first simulation (Imagine vs implicit baseline) across both groups including (see Table 3S): (1) right and (2) left temporal cortex (spanning the middle temporal gyrus, Heschl gyrus, angular gyrus, and areas of the inferior frontal gyrus (IFG), pars opercularis), (3) cerebellum and fusiform occipital cortex, (4) posterior cingulate and supplementary motor area (SMA), (5) left temporal pole, and (6) left hippocampus, Figure 1Sa. Two bilateral clusters showed significant parametric activation associated with higher unpleasantness ratings during the first simulation (Imagine vs implicit baseline) across both groups, including: insula, temporal pole, opercular cortex and IFG pars opercularis, Figure 1Sb.

There were no between group differences in any of the above brain areas, whose activation was parametrically modulated by subjective ratings of realness and unpleasantness the Imagine condition.

#### Resting state

There was no significant difference in RSN functional connectivity before and after the future simulation task across the whole sample (main effect of time). There was also no significant Time x Group interaction. However, there was a main effect of Group (both resting state periods combined, Table 3), showing that compared to HCs the BPDS group had: 1) greater intrinsic RSN functional connectivity within the default mode network, in a cluster located across the right MFG and IFG; and 2) greater RSN functional connectivity between the sensori-motor (SM) network and a cluster located in the right frontal pole, paracingulate gyrus and superior frontal gyrus (SFG).

**Table 3.**
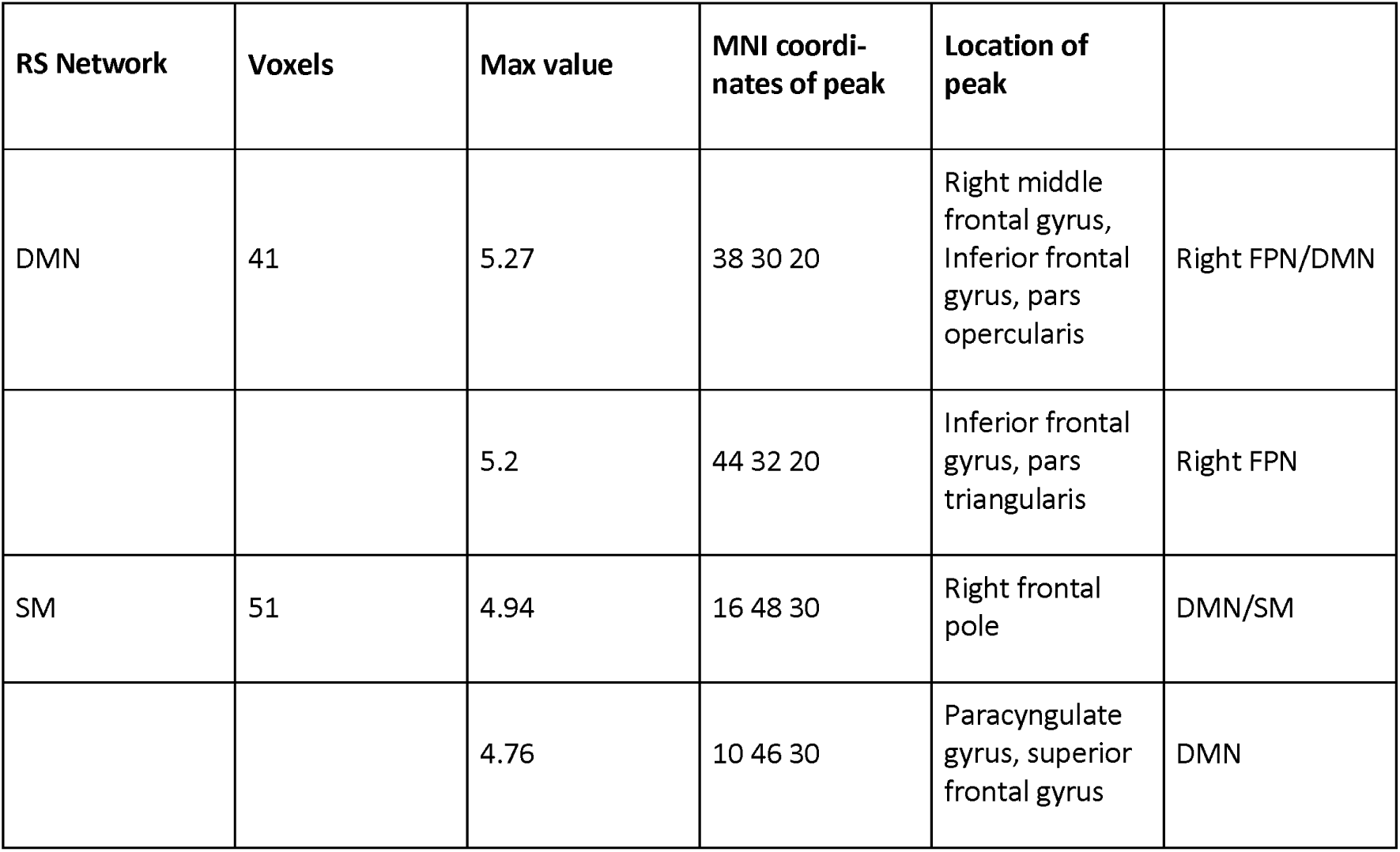
Clusters showing significant differences in RSN connectivity comparing the BPSD to the HC group across both resting-state acquisitions.

## 4. Discussion

### Summary of findings

To our knowledge, this study is the first to investigate the neural bases of mental imagery-based episodic simulation of negative future scenarios (negative episodic future simulation) in BPSD, as well as the subjective and neural impact of perceptual modifications of such future imagery.

As hypothesised, BPDS individuals reported greater subjective perceptual realness and emotional unpleasantness of their negative episodic future simulation than did healthy controls (HCs), which produced a transient increase in state sadness in the BPSD group only. Critically and as predicted, perceptual modification of negative episodic future simulation (i.e. turning the mental images from colour to black and white, relative to a control of simple repetition), successfully reduced realness and unpleasantness ratings in BPDS individuals to levels equivalent to HC individuals.

Across both groups, subjective perceptual realness of episodic future imagery was associated with neural activity in areas comprising the so-called ‘core network’ of episodic memory and episodic future simulation^21^, while subjective unpleasantness of the imagery was associated with activity in the insula and surrounding areas.

At the neural level, the BPDS group exhibited greater neural activity during the initial negative episodic future simulation compared to healthy controls in PFC areas of frontal pole, MFG, IFG, orbitofrontal cortex and insula, pointing to a dysfunctional core simulation circuitry in BPDS. No between-group differences in neural activity were found during perceptual modification of negative episodic future simulation. Relative to healthy controls, BPDS individuals also showed greater RSN functional connectivity (before and after simulation) in regions of the DMN.

### Subjective perceptual characteristics and emotional impact of episodic future simulation and its perceptual modification

In keeping with previous findings^9,14–16^, our study provides additional evidence that negative episodic future simulation has a greater emotional impact on BPDS individuals than healthy controls, even during euthymic states. Importantly, significant increases in sadness in BPDS after imagining common negative daily events inside an artificial MRI scanner environment supports the hypothesised emotional amplification role of emotional future mental imagery in this population^35^.

We tested for the first time a laboratory analogue of a clinically relevant mental imagery perceptual modification technique^6,7^. As individuals with BPDS describe that their mental images feel “as if they were real” and often make them act accordingly, the perceptual modification of imagery realness (e.g. shrinking images, turning them into black and white) during therapy reduces perceived imagery realness without engaging with the imagery’s content meaning^8^. This can be advantageous when imagery has variable contents or when patients avoid its distressing content. Using a controlled experimental task, the present study demonstrates that BPDS individuals can easily engage with such perceptual modification techniques, and this intervention was able to reduce both the perceptual characteristics and emotional impact of such imagery to healthy control levels.

There is growing recognition that understanding distortions in episodic future simulation can inform experimental-based intervention development for mood and anxiety disorders^9^. We highlight the need to further investigate the therapeutic potential of simple perceptual modification techniques targeting imagery in BPSD, which can readily be combined with pharmacological approaches and may be easier for patients who struggle with traditional (rational) thought-challenging in verbal cognitive-behavioural therapy.

### Neural bases of deliberate future simulation of negative scenarios

Importantly, BPDS individuals showed greater neural activity during episodic future simulation in lateral PFC networks regulating social, emotional and planning aspects of future simulation^36^ that have been previously reported as key dysfunctional brain networks in bipolar disorder. Research has found both structural and functional abnormalities in the IFG^37,38^, altered connectivity within MFG, IFG and frontal pole areas^37,38^ and altered functional connectivity within the fronto-insular network^39^ in BPSD compared to controls in relation to emotion processing and regulatory control.

In relation to future simulation, these areas are at the interface of the default mode network (DMN), frontoparietal and frontoinsular networks of the brain supporting flexible and adaptive self-generated thought^40^. Frontopolar regions activate when orienting one’s internal attention to future events that include selfand social awareness^41^. Insula/IFG activity is linked to physiological arousal and social influence on behaviour^42^. The frontoparietal network helps control self-generated emotional internal thought^20^ and planning^43^. Instead, our ROI analysis did not find evidence of abnormal activity in the amygdala in areas related to vividness or emotional valence of simulation in BDSD compared to controls. Hence, our data suggest that a mechanism contributing to emotional dysregulation in BPSD may be higher attention orienting^42^ and higher salience attribution to future imagery including its social and somatic arousal aspects^69^. In turn, this may trigger greater cognitive control effort^44^.

In keeping with a spectrum approach to psychopathology, functional impairment in the IFG is also present in individuals with BPDS based on high genetic risk^45^ and may indicate specific bipolar disorder risk in young people with transdiagnostic emotional dysregulation^46^. Recent evidence highlights ventrolateral PFC areas such as IFG/frontal pole as key for BPSD^47^, as they signal dysfunction in reward expectancy associated with impulsivity and frustration. Our data bring additional evidence that in BPSD, increased activity in lateral frontopolar areas involved in forecasting and maintaining information relevant to future decisions^48^ is associated with episodic future simulation. Further studies should test whether this also applies to simulation of potentially rewarding scenarios. Importantly, future simulation may represent a key cognitive process via which forecasting dysfunction manifests phenomenologically and translates into the patients’ experience of future mental imagery as compelling and driving behaviour^49,50^. The frontoparietal network’s role in scene construction and planning^20,43^ further supports the relevance of simulation for influencing action in this group.

Overall our findings are the first to demonstrate that negative future simulation is underpinned by altered activity in key cortical hubs associated to impaired emotional regulation in BPDS, providing neurofunctional evidence for the role of emotional mental imagery and in particular future simulation in BPDS emotional dysregulation. We propose that the role of future simulation in BPDS psychopathology may be linked to maladaptive content aspects of the imagery (social and somatic salience), dysfunctional control over the simulation in relation to the attention it receives and possible impact on planning behaviour, rather than to heightened emotional reactivity to internal imaginal stimuli.

### Resting state functional connectivity following future simulation

Our study also detected RSN functional connectivity differences between euthymic BPDS individuals and controls within a small right lateral PFC area part of the DMN at the interface with the frontoparietal network. If replicated, this contrasts with a recent systematic review ^18^ concluding that based on ICA methodology, euthymic bipolar patients do not show RSN functional connectivity alterations *within* the DMN, frontoparietal or salience networks, possibly indicating a state of illness remission. The discrepancy may be explained by the fact that, unique to our study, we measured RSN functional connectivity after an episodic future simulation task. However, we found no direct evidence of carry-over effect of imagery during rest (i.e. spontaneously occurring future simulation).

### Limitations

Due to image acquisition problems and motion artefacts (see Supplementary Material) our initial sample size was reduced from N=25 and N=23 to N=20 and N=17. Moreover, our main findings were from an exploratory whole-brain analysis. This necessitates replication in larger samples that also allow controlling for medication effects. Additionally, the small cluster size of our RSN findings warrants further testing. While we chose a spectrum approach, based on imagery and emotion dysregulation evidence in at risk groups^22,23,82–84^, our findings should also be replicated in purely clinical samples to extrapolate definitive implications for the neurobiology and treatment of bipolar disorder. Moreover, our study design cannot distinguish what might be a bipolarity trait marker from a neural marker of disease. Another limitation is that despite our BPDS group being euthymic based on the SCID, it still presented higher subthreshold low mood and anxiety compared to HC, which may confound findings on the cognitive process of episodic future simulation. However, we argue that complete absence of low mood or anxiety would not represent BPDS phenomenology. Finally, we did not acquire a measure of mood instability or emotional dysregulation.

## 5. Conclusions

We argue that dysfunctional mental imagery-based simulations of emotional future events can contribute to emotional dysregulation in BPDS. Our study supports the growing evidence that individuals with BPDS experience more emotionally intense mental imagery of negative future events than do healthy individuals, even during euthymic states. Critically, this is underpinned by aberrant future simulation circuitry across the default mode network, frontoparietal and fronto-insular areas, suggesting that heightened perpetual and emotional strength of future imagery in BPSD individuals reflect aberrant control over simulation. We propose that dysfunctional regulatory circuitry of the social, emotional and planning functions of simulation is a mechanism via which mental imagery amplifies emotion in BPDS. Our findings also provide preliminary indication that episodic future simulation may be an important cognitive process bridging brain dysfunction during emotion regulation and resting state in bipolar disorder. Its role in mood instability warrants further investigation. Importantly, in keeping with emerging clinical practice, the present results provide evidence that anomalies in future imagery can be a tractable target for intervention, and simple perceptual modifications of such imagery could be a promising emotion regulation technique for BPDS.

## Data Availability

All data produced in the present study are available upon reasonable request to the authors.

## Acknowledgements

We would like to thank all participants in the study. We would like to thank Peter Watson from the MRC Cognition and Brain Sciences Unit, University of Cambridge for his support with the statistical analysis, and Russell Thompson from the MRC Cognition and Brain Sciences Unit, University of CambridgeU for support with neuroimaging data processing and analysis.

## REFERENCES

1. Ferrari AJ. The prevalence and burden of bipolar disorder: findings from the Global Burden of Disease Study 2013. Bipolar Disorders. 2016;18:440–450.

2. Merikangas KR, Jin R, He JP, et al. Prevalence and correlates of bipolar spectrum disorder in the world mental health survey initiative. Arch Gen Psychiatry. Mar 2011;68(3):241–51. doi:10.1001/archgenpsychiatry.2011.12

3. Bopp JM, Miklowitz DJ, Goodwin GM, Stevens W, Rendell JM, Geddes JR. The longitudinal course of bipolar disorder as revealed through weekly text messaging: a feasibility study. Bipolar Disord. May 2010;12(3):327–34. doi:10.1111/j.1399-5618.2010.00807.x

4. Harrison PJ, Cipriani A, Harmer CJ, et al. Innovative approaches to bipolar disorder and its treatment. Ann N Y Acad Sci. Feb 2016;1366(1):76–89. doi:10.1111/nyas.13048

5. Goodwin GM, Haddad PM, Ferrier IN, et al. Evidence-based guidelines for treating bipolar disorder: Revised third edition recommendations from the British Association for Psychopharmacology. J Psychopharmacol. Jun 2016;30(6):495–553. doi:10.1177/0269881116636545

6. Holmes EA, Bonsall MB, Hales SA, et al. Applications of time-series analysis to mood fluctuations in bipolar disorder to promote treatment innovation: a case series. Transl Psychiatry. Jan 26 2016;6:e720. doi:10.1038/tp.2015.207

7. Holmes EA, Hales, S. A., Young, K. & Di Simplicio, M. magery-Based Cognitive Therapy for Bipolar Disorder and Mood Instability. Guildford Press; 2019.

8. Pine DS, Wise SP, Murray EA. Evolution, Emotion, and Episodic Engagement. Am J Psychiatry. Aug 1 2021;178(8):701–714. doi:10.1176/appi.ajp.2020.20081187

9. Di Simplicio M, Renner F, Blackwell SE, et al. An investigation of mental imagery in bipolar disorder: Exploring “the mind’s eye”. Bipolar Disord. Dec 2016;18(8):669–683. doi:10.1111/bdi.12453

10. Pearson J, Naselaris T, Holmes EA, Kosslyn SM. Mental Imagery: Functional Mechanisms and Clinical Applications. Trends Cogn Sci. Oct 2015;19(10):590–602. doi:10.1016/j.tics.2015.08.003

11. Ji JL, Heyes SB, MacLeod C, Holmes EA. Emotional Mental Imagery as Simulation of Reality: Fear and Beyond-A Tribute to Peter Lang. Behav Ther. Sep 2016;47(5):702–719. doi:10.1016/j.beth.2015.11.004

12. Ji JL, Kavanagh DJ, Holmes EA, MacLeod C, Di Simplicio M. Mental imagery in psychiatry: conceptual & clinical implications. CNS Spectr. Feb 2019;24(1):114–126. doi:10.1017/S1092852918001487

13. Ji JL, Geiles D, Saulsman LM. Mental imagery-based episodic simulation amplifies motivation and behavioural engagement in planned reward activities. Behav Res Ther. Oct 2021;145:103947. doi:10.1016/j.brat.2021.103947

14. Di Simplicio M, Lau-Zhu A, Meluken I, et al. Emotional Mental Imagery Abnormalities in Monozygotic Twins With, at High-Risk of, and Without Affective Disorders: Present in Affected Twins in Remission but Absent in High-Risk Twins. Front Psychiatry. 2019;10:801. doi:10.3389/fpsyt.2019.00801

15. Holmes EA, Deeprose C, Fairburn CG, et al. Mood stability versus mood instability in bipolar disorder: a possible role for emotional mental imagery. Behav Res Ther. Oct 2011;49(10):707–13. doi:10.1016/j.brat.2011.06.008

16. O’Donnell C, Di Simplicio M, Brown R, Holmes EA, Burnett Heyes S. The role of mental imagery in mood amplification: An investigation across subclinical features of bipolar disorders. Cortex. Aug 2018;105:104–117. doi:10.1016/j.cortex.2017.08.010

17. Phillips ML, Swartz HA. A critical appraisal of neuroimaging studies of bipolar disorder: toward a new conceptualization of underlying neural circuitry and a road map for future research. Am J Psychiatry. Aug 2014;171(8):829–43. doi:10.1176/appi.ajp.2014.13081008

18. Syan SK, Smith M, Frey BN, et al. Resting-state functional connectivity in individuals with bipolar disorder during clinical remission: a systematic review. J Psychiatry Neurosci. Aug 2018;43(5):298–316.

19. Wang Y, Gao Y, Tang S, et al. Large-scale network dysfunction in the acute state compared to the remitted state of bipolar disorder: A meta-analysis of resting-state functional connectivity. EBioMedicine. Apr 2020;54:102742. doi:10.1016/j.ebiom.2020.102742

20. Andrews-Hanna JR, Smallwood J, Spreng RN. The default network and self-generated thought: component processes, dynamic control, and clinical relevance. Ann N Y Acad Sci. May 2014;1316:29–52. doi:10.1111/nyas.12360

21. Benoit RG, Schacter DL. Specifying the core network supporting episodic simulation and episodic memory by activation likelihood estimation. Neuropsychologia. Aug 2015;75:450–7. doi:10.1016/j.neuropsychologia.2015.06.034

22. Hirschfeld RM. The Mood Disorder Questionnaire: A Simple, Patient-Rated Screening Instrument for Bipolar Disorder. Prim Care Companion J Clin Psychiatry. Feb 2002;4(1):9–11. doi:10.4088/pcc.v04n0104

23. First M, Spitzer, R., Gibbon, M. & Williams, J. Structured Clinical Interview for DSM-IV Axis I Disorders. Biometrics Research; 2002.

24. Rush AJ, Trivedi MH, Ibrahim HM, et al. The 16-Item Quick Inventory of Depressive Symptomatology (QIDS), clinician rating (QIDS-C), and self-report (QIDS-SR): a psychometric evaluation in patients with chronic major depression. Biol Psychiatry . Sep 1 2003;54(5):573–83. doi:10.1016/s0006-3223(02)01866-8

25. Altman EG, Hedeker D, Peterson JL, Davis JM. The Altman Self-Rating Mania Scale. Biol Psychiatry. Nov 15 1997;42(10):948–55. doi:10.1016/S0006-3223(96)00548-3

26. Nelis S, Holmes EA, Griffith JW, Raes F. Mental imagery during daily life: Psychometric evaluation of the Spontaneous Use of Imagery Scale (SUIS). Psychol Belg. Jan 20 2014;54(1):19–32. doi:10.5334/pb.ag

27. O’Donnell C, Di Simplicio M, Burnett Heyes S. Hypomanic-like experiences and spontaneous emotional mental imagery. J Affect Disord. Dec 1 2020;277:742–746. doi:10.1016/j.jad.2020.08.003

28. Marks DF. Visual imagery differences in the recall of pictures. Br J Psychol. Feb 1973;64(1):17–24. doi:10.1111/j.2044-8295.1973.tb01322.x

29. Deeprose C, Holmes EA. An exploration of prospective imagery: the impact of future events scale. Behav Cogn Psychother. Mar 2010;38(2):201–9. doi:10.1017/S1352465809990671

30. Beck AT, Epstein N, Brown G, Steer RA. An inventory for measuring clinical anxiety: psychometric properties. J Consult Clin Psychol. Dec 1988;56(6):893–7. doi:10.1037//0022-006x.56.6.893

31. Henry C, M’Bailara K, Lepine JP, Lajnef M, Leboyer M. Defining bipolar mood states with quantitative measurement of inhibition/activation and emotional reactivity. J Affect Disord. Dec 2010;127(1-3):300–4. doi:10.1016/j.jad.2010.04.028

32. Smith SM, Jenkinson M, Woolrich MW, et al. Advances in functional and structural MR image analysis and implementation as FSL. Neuroimage. 2004;23 Suppl 1:S208–19. doi:10.1016/j.neuroimage.2004.07.051

33. Woolrich MW, Behrens TE, Beckmann CF, Jenkinson M, Smith SM. Multilevel linear modelling for FMRI group analysis using Bayesian inference. Neuroimage. Apr 2004;21(4):1732–47. doi:10.1016/j.neuroimage.2003.12.023

34. Beckmann CF, DeLuca M, Devlin JT, Smith SM. Investigations into resting-state connectivity using independent component analysis. Philos Trans R Soc Lond B Biol Sci. May 29 2005;360(1457):1001-13. doi:10.1098/rstb.2005.1634

35. Holmes EA, Geddes JR, Colom F, Goodwin GM. Mental imagery as an emotional amplifier: application to bipolar disorder. Behav Res Ther. Dec 2008;46(12):1251–8. doi:10.1016/j.brat.2008.09.005

36. Hassabis D, Maguire EA. The construction system of the brain. Philos Trans R Soc Lond B Biol Sci. May 12 2009;364(1521):1263-71. doi:10.1098/rstb.2008.0296

37. Roberts G, Lord A, Frankland A, et al. Functional Dysconnection of the Inferior Frontal Gyrus in Young People With Bipolar Disorder or at Genetic High Risk. Biol Psychiatry. Apr 15 2017;81(8):718–727. doi:10.1016/j.biopsych.2016.08.018

38. Anand A, Grandhi J, Karne H, Spielberg JM. Intrinsic functional connectivity during continuous maintenance and suppression of emotion in bipolar disorder. Brain Imaging Behav. Oct 2020;14(5):1747–1757. doi:10.1007/s11682-019-00109-4

39. Ellard KK, Gosai AK, Felicione JM, et al. Deficits in frontoparietal activation and anterior insula functional connectivity during regulation of cognitive-affective interference in bipolar disorder. Bipolar Disord. May 2019;21(3):244–258. doi:10.1111/bdi.12709

40. Golchert J, Smallwood J, Jefferies E, et al. Individual variation in intentionality in the mind-wandering state is reflected in the integration of the default-mode, fronto-parietal, and limbic networks. Neuroimage. Feb 1 2017;146:226–235. doi:10.1016/j.neuroimage.2016.11.025

41. Okuda J, Fujii T, Ohtake H, et al. Thinking of the future and past: the roles of the frontal pole and the medial temporal lobes. Neuroimage. Aug 2003;19(4):1369–80. doi:10.1016/s1053-8119(03)00179-4

42. Menon V, Uddin LQ. Saliency, switching, attention and control: a network model of insula function. Brain Struct Funct. Jun 2010;214(5-6):655–67. doi:10.1007/s00429-010-0262-0

43. Spreng RN, Stevens WD, Chamberlain JP, Gilmore AW, Schacter DL. Default network activity, coupled with the frontoparietal control network, supports goal-directed cognition. Neuroimage. Oct 15 2010;53(1):303–17. doi:10.1016/j.neuroimage.2010.06.016

44. Hampshire A, Chamberlain SR, Monti MM, Duncan J, Owen AM. The role of the right inferior frontal gyrus: inhibition and attentional control. Neuroimage. Apr 15 2010;50(3):1313–9. doi:10.1016/j.neuroimage.2009.12.109

45. Breakspear M, Roberts G, Green MJ, et al. Network dysfunction of emotional and cognitive processes in those at genetic risk of bipolar disorder. Brain. Nov 2015;138(Pt 11):3427–39. doi:10.1093/brain/awv261

46. de Oliveira L, Portugal LCL, Pereira M, et al. Predicting Bipolar Disorder Risk Factors in Distressed Young Adults From Patterns of Brain Activation to Reward: A Machine Learning Approach. Biol Psychiatry Cogn Neurosci Neuroimaging. Aug 2019;4(8):726–733. doi:10.1016/j.bpsc.2019.04.005

47. Edmiston EK, Fournier JC, Chase HW, et al. Assessing Relationships Among Impulsive Sensation Seeking, Reward Circuitry Activity, and Risk for Psychopathology: A Functional Magnetic Resonance Imaging Replication and Extension Study. Biol Psychiatry Cogn Neurosci Neuroimaging. Jul 2020;5(7):660–668. doi:10.1016/j.bpsc.2019.10.012

48. Boorman ED, Behrens TE, Rushworth MF. Counterfactual choice and learning in a neural network centered on human lateral frontopolar cortex. PLoS Biol. Jun 2011;9(6):e1001093. doi:10.1371/journal.pbio.1001093

49. Hales SA, Deeprose C, Goodwin GM, Holmes EA. Cognitions in bipolar affective disorder and unipolar depression: imagining suicide. Bipolar Disord. Nov-Dec 2011;13(7-8):651–61. doi:10.1111/j.1399-5618.2011.00954.x

50. Ivins A, Di Simplicio M, Close H, Goodwin GM, Holmes E. Mental imagery in bipolar affective disorder versus unipolar depression: investigating cognitions at times of ‘positive’ mood. J Affect Disord. Sep 2014;166:234–42. doi:10.1016/j.jad.2014.05.007

