## Supplementary Material for "Neural basis of negative future mental imagery-based simulation in the bipolar disorder spectrum"

**2.1 Participants and procedures**

*Details of screening and exclusion criteria*

Potentially eligible participants were invited for the screening session if they were a) eligible for MRI scanning, and b) scored either below 3 or above 7 on the MDQ. Fifty-five such individuals were administered the SCID^1^ by a trained psychiatrist during the screening session to assess lifetime psychiatric history. Participants were invited to the study if they reported a) an absence of a current mood episode (i.e., were euthymic). Two participants were excluded, one based on current depressive episode, and one for not meeting criteria for the bipolar disorder spectrum group as described in the study protocol. The final sample comprised of *N*=26 participants with BPSD and 27 healthy controls matched on age and gender.

After screening, eligible participants completed two separate experimental sessions: a behavioural testing session including computer-based tasks (not reported in this paper) and pen and paper questionnaires, and a neuroimaging session comprising the acquisition of structural MRI data followed by functional MRI (fMRI) scanning during a future simulation task and during rest. Four participants did not complete the neuroimaging session due to the following reasons: MRI incompatibility not disclosed at pre-screening (*n* = 1); scan interrupted due to paresthesias or headache (*n* = 2); drop out (*n* = 1). Of the remaining 49 participants, data were excluded from 5 participants due to MRI acquisition problems (‘ghosting’, misplaced scan box), and from 1 participant due to structural brain abnormalities. This resulted in a final sample of 43 participants, from which a further 6 were excluded due to excessive motion artefacts (>2 mm relative displacement in any direction). Results are reported for the remaining *n* = 20 participants in the BPSD group and *n* = 17 in the HC group.

**2.2 Measures and task**

*2.2.1. Questionnaires*

Participants completed the following questionnaires assessing mental imagery-based cognition.

*Baseline mental imagery tendency and ability*

The *Spontaneous Use of Imagery Scale* (SUIS)^2^ is a 12-items scale assessing the general tendency to use mental imagery in everyday life (e.g., “When I think about a series of errands I must do, I visualize the stores I will visit”). Participants indicate the degree to which each sentence is appropriate for them on a 5-point Likert scale (1 = “never appropriate; 5 = “completely appropriate”). The SUIS has shown good reliability (Cronbach’s α = 0.74; test-retest: *r* = .53) and is moderately correlated with measures of visual imagery ability, such as the Vividness of Visual Imagery Questionnaire (*r* = 0.38) 3 and the Plymouth Sensory Imagery Questionnaire (*r* = 0.33) 4, and lowly correlated with non-visual imagery ability, such as taste imagery (*r* = 0.18) (41), indicating good construct validity.

The *Spontaneous Use of Emotional Mental Imagery Scale* (E-SUIS)^5^ is a 16 items scale adapted from the SUIS assessing the tendency to general use of mental imagery during daily life in association with emotional events (e.g. “I’m going on a trip to somewhere I haven’t gone before and I’m very excited about. I visualize what the place will be like”). Participants indicate the degree to which each sentence is appropriate for them on a 5-point Likert scale (1 = “never appropriate, 5 = “completely appropriate”). The E-SUIS has shown good internal consistency (Cronbach’s Alpha =.88) and in a sample of young adults across levels of hypomanic experiences 5.

The *Vividness of Visual Imagery Questionnaire* (VVIQ)^3^ is a 16-item scale assessing the ability to generate vivid mental imagery. Individuals are asked to imagine four daily life scenarios unfolding (e.g., “Visualise the rising sun”), each guided by four descriptive sentences (e.g., from the sun rising, to the sky clearing, to a storm gathering, to a rainbow appearing). The vividness of imagery cued by each sentence is rated on a 5-point Likert scale (1 = “perfectly clear and as vivid as normal vision”, 5 = “No image at all, you only know you are thinking of the object”). The VVIQ has shown acceptable internal consistency (Cronbach’s alpha= .76).

The *Plymouth Sensory Imagery Questionnaire* (Psi-Q)^4^ is a 35-item scale assessing the ability to generate the vividness of mental imagery across seven modalities (visual, auditory, olfactory, taste, touch, bodily sensation, and emotional feelings). Individual are asked to imagine and rate the vividness of five scenes per category (e.g. visual “a bon fire”; auditory: “the mewing of a cat”; smell: “fresh paint”) on an 11-point Likert scale (0 = no image at all, 10 = image as clear and vivid as real life). The Psi-Q has shown good reliability (Cronbach’s α = .97 ; test-retest: *r* = 0.71), and moderate to high correlations with measures of imagery ability such as the VVIQ (*r* = 0.66), and the visual subscale (more so than non-visual subscales) show low to moderate correlations with measures of visual imagery tendency, such as the SUIS (*r* = 0.33) (41), indicating good construct validity 4.

The *Impact of Future Events Scale* (IFES)^6^ is a 24 items scale assessing the impact of imagining future events. Individuals are first asked to report three future events that they have been imagining over the last seven days, and to indicate whether each event was “positive” or “negative”. Next, participants rated a series of statements describing the impact of imagining future events in the past week on them, relating to intrusive pre-experiencing, avoidance and hyper-arousal domains (e.g., “I stayed away from reminders of the future”). Each statement was rated on a 5-point Likert scale of how true it was for the participant, ranging from 0 = Not at all to 4 = Extremely. The IFES has shown good reliability (Cronbach’s α = 0.87; test-retest: *r* = .73).

*Affective psychopathology*

The *Altman Self-Rating Scale for Mania* (ASRM)^7^ is a widely used 5-item scale assessing the frequency and severity of manic symptoms on a range from 0 to 4 over the past 7 days (e.g., 0 = I do not feel more self-confident than usual; 4 = I feel extremely self-confident all of the time). Total ASRM scores range from 0 to 20, where scores > 6 indicate significant manic or hypomanic symptoms. The ASRM has established psychometric properties for detecting mania in patients with BD.

The *Quick Inventory of Depressive Symptomatology, Self-Report* (QIDS-SR)^8^ is a 16-item questionnaire measuring the severity of depression over the past 7 days on a 4-point scale (0–3) anchored at all points by a description (e.g., Feeling Sad: 0 = I do not feel sad; 3 = I feel sad nearly all of the time). The QIDS-SR has established psychometric properties for rating depressive symptom severity in individuals with major depression. Scores correspond to five levels of depression severity: mild = 6–10; moderate = 11–15; severe = 16–20; very severe = 21–27. Scores < 6 typically indicate euthymia.

The *Beck Anxiety Inventory* (BAI)^9^ is a 21 items scale assessing common symptoms of anxiety (e.g., numbness and tingling, sweating not due to heat, fear of the worst happening etc.) over the past 7 days, on a 4-point scale (0 = Not at all; 3 = Severely – it bothered me a lot). Scores range from 0-7 = minimal anxiety, 8-15 = mild anxiety, 16-25 = moderate anxiety and 26-63 = severe anxiety. It has well established psychometric properties and has been shown to measure clinical anxiety with minimal overlap with depressive symptoms.

The *Multidimensional Assessment of Thymic States* (MAThyS)^10^ is a 20 items scale assessing emotional reactivity over the last 7 days. The items cover five dimensions: emotional reactivity, cognition speed, psychomotor function, motivation and sensory perception. The second part of the test evaluates the frequency with which the participant has experienced seven emotions during the previous week, on a five-point Likert scale running from "never" to "constantly." The emotions evaluated are sadness, joy, irritability, panic, anxiety, anger and euphoria. Individuals with BPDSD with mixed feature are supposed to have high scores in this scale in terms of “intensity”, in comparison with depressed individuals which are often described as insensitive or anhedonic. Confirmatory analyses demonstrated a good validity for the dimensions of motivation and psychomotor function, and a good internal consistency (Cronbach’s. Alpha =.95).

*2.2.2. Episodic Future Simulation Task*

The Episodic Future Simulation Task involved reading and imagining 20 mildly negative scenarios describing common daily situations, half of which involved interpersonal situations, based on^11^. To maximise scenario realism, each scenario was presented in a unfolding episodic manner via four gradually appearing sentences written in present tense, with the last sentence describing physiological responses to the situation without affective labels, e.g.: “It's Monday morning and you are in bed/ Your alarm goes off and you hit the snooze button/ Seeing how late it is, you almost leap out of bed/ Realising you will be late for a key meeting, your heart is pounding”. Participants were instructed to read the sentences whilst building an unfolding episodic future event in their mind, to visualise it as if seeing from their own eyes, as if the situation was really happening in a familiar place and with familiar people.

**2.3 FMRI procedure and data acquisition**

MRI data were acquired on a 3T Siemens TIM Trio System scanner at the MRC CBSU, Cambridge, using a 32-channel head coil. A high–resolution 3D whole-brain T1-weighted structural image was acquired for registration purposes, using an MP RAGE sequence, with voxel resolution of 1 x 1 x 1 mm^3^, echo time = 3.02 ms, and repetition time = 2250 ms. Functional scans were obtained using an EPI-BOLD contrast image with a total of 32 slices, with a voxel resolution of 3 x 3 x 3 mm^3^, repetition time = 2000 ms, echo time = 30 msec, flip angle = 78°.

2.4 Analysis

*2.4.3.1. FMRI data: episodic future simulation task*

*Data preprocessing*

For the future simulation task data, pre-processing consisted of head motion correction (using MCFLIRT)^12^ and brain extraction (using FSL’s Brain Extraction Tool, BET)^13^. Then, each individual data were further processed by applying the FSL Independent Component Analysis tool to each individual subject separately^14^ to remove artefacts and separate neural-related signal from different sources of noise. Following the guidelines for hand classification of fMRI ICA noise components by^15^, components that represented obvious scanner-related or physiological artefacts were identified and removed from each subject’s individual data. Two experimenters (MDS, RMV) independently conducted this procedure on 10% of data (5 participants); after establishing a 91.5% inter-rater reliability at classifying components as noise or signal, one experimenter (MDS) completed the hand classification on all individual level data and ambiguous data (average of 13.8% of components per individual) were double-checked by the second experimenter (RMV) until consensus was reached. Further data processing used FEAT (FMRI Expert Analysis Tool) Version 6.00, and included spatial smoothing using a Gaussian kernel of FWHM (full width at half maximum) 5mm, intensity normalisation and highpass temporal filtering set at 90s. Functional data were registered to high resolution images and then to standard-space using FSL’s BBR and non-linear registration tools^16,17^.

*2.4.3.1. FMRI data: resting state*

*Data preprocessing*

For resting state (RS) functional data pre-processing involved brain extraction using the Brain Extraction Tool 13, motion correction using MCFLIRT^12^, spatial smoothing using a Gaussian kernel of 6mm FWHM, grand-mean intensity normalisation of the entire 4D dataset by a single multiplicative factor, and highpass temporal filtering with a cutoff of 150s. Functional data was registered to each participant’s structural image using FMRIB’s Linear Image Registration Tool (FLIRT)^17^ and optimised using the Boundary-Based Registration (BBR) technique^16^. Images were then registered to standard space (MNI-152 template) using FMRIB’s Nonlinear Registration Tool (FNIRT)^18^.

Independent components analysis (ICA) was conducted using the MELODIC tool. A single-session ICA design was used to decompose each individual’s functional data into components. FMRIB’s ICA-based Xnoiseifier (FIX)^19^, was used to auto-classify components as signal or noise, based on hand-classified training data from a separate dataset. Components defined as noise were removed, and the subsequent denoised data was then registered to standard space as above.

Other relevant references for fMRI data analysis procedures: ^20-22^

**Results**

**Table 1S. Clinical characteristics of the BPDS group**

| **Diagnosis** | **BPSD (n=20)** |
| --- | --- |
| Bipolar I, n | 6 |
| Bipolar II, n | 3 |
| BP NOS, n | 7 |
| High spectrum (MDQ >7), n | 4 |
| Age of onset M (SD) | 14.6 (3.3) |
| **History of Depression,** N of episodes, n |  |
| 0 | 1 |
| 1 - 4 | 9 |
| 5 – 9 | 5 |
| >= 10 | 5 |
| **History of Mania,** N of episodes, n |  |
| 0 | 7 |
| 1 - 4 | 5 |
| 5 – 9 | 4 |
| >= 10 | 4 |
| **History of suicide attempts, n** | 4 |
| **Current Anxiety Disorder comorbidity, n (%)** | 10 (50) |
| Social anxiety, n | 3 |
| Obsessive compulsive disorder, n | 1 |
| Panic Disorder / Agoraphobia, n | 2 |
| Post-traumatic stress disorder, n | 2 |
| Generalized anxiety disorder, n | 1 |
| Specific phobia, n | 3 |
| **Anxiety disorder lifetime comorbidity, n (%)** | 15 (78.9) |
| **Current Other Axis I comorbidity, n (%)** | 1 (5) |
| Eating Disorder Substance dependence, n | 1 |
| **Medication** |  |
| Antidepressant, n | 4 |
| Mood stabilizer, n | 5 |

**3.3 Behavioural data: Episodic Future Simulation Task**

| **Table 2S.**  Episodic future simulation task: imagery ratings | | | |
| --- | --- | --- | --- |
|  | **BPSD (n=20)**  **Mean (SD)** | **Healthy Controls (n=17)**  **Mean (SD)** | ***Z*** |
| ***Realness of imagined scenario (Likert scale 0-10)*** | | | |
| Imagine | 7.1 (1.5) | 5.9 (1.8) | Z=2.71, p=.007 |
| Change to Black & White | 5.9 (2.2) | 5.1 (1.8) | Z=1.15, p=.25 |
| Repeat | 6.9 (1.8) | 5.7 (2) | Z=2.34, p=.018 |
| ***Unpleasantness of imagined scenario (Likert scale 0-10)*** | | | |
| Imagine | 6.3 (1.5) | 5.1 (1.7) | Z=2.61, p=.009 |
| Change to Black & White | 5.2 (1.7) | 4.3 (1.8) | Z=1.68, p=.09 |
| Repeat | 6.1 (1.9) | 5.1 (1.9) | Z=1.90, p=.057 |

**3.4 FMRI data**

Brain-behaviour correlations

**Table 3S.**

Clusters showing significant functional activation during episodic future simulation (Imagine condition) associated with subjective ratings of simulation realness and unpleasantness across the whole sample.

| **Contrast** | **Voxels** | **P value / Z** | **MNI coordinates of peak** | **Location of peak** |
| --- | --- | --- | --- | --- |
| Positive correlation with realness ratings during Imagine | 4810 | 5.96e-08 / 5.02 | 64 -4 4 | Right temporal cortex (middle temporal gyrus, Heschl gyrus, angular gyrus, and IFG, pars opercularis). Figure 3 |
|  | 1648 | 0.001/4.54 | -20 -56 -22 | Left temporal cortex (middle temporal gyrus, Heschl gyrus, angular gyrus, and IFG, pars opercularis) |
|  | 1363 | 0.005/3.77 | 4 -16 44 | Right cerebellum and fusiform occipital cortex |
|  | 1329 | 0.006/4.36 | -58 0 -12 | Posterior cingulate and supplementary motor area |
|  | 916 | 0.041 /4.1 | -56 -24 8 | Left temporal pole |
|  | 884 | 0.048 / 4.13 | -56 30 14 | Left hippocampus |
| Positive correlation with unpleasantness during Imagine | 1575 | 0.002 / 3.68 | 56 12 -6 | Right insula, temporal pole, opercular cortex and IFG pars opercularis. |
|  | 941 | 0.040 / 3.48 | -58 0 -12 | Left insula, temporal pole, opercular cortex and IFG pars opercularis. |

**Figure 1S. Neural activity of the core simulation network during episodic future imagery across the whole sample.**

Areas of parametric activation in core simulation network as a function of realness ratings (A) and unpleasantness ratings (B) during future simulation (Imagine) across both groups. Activation is thresholded using clusters determined by Z>2.3 (P<0.01) and a (corrected) cluster significance threshold of P=0.05. L = Left, R = Right.

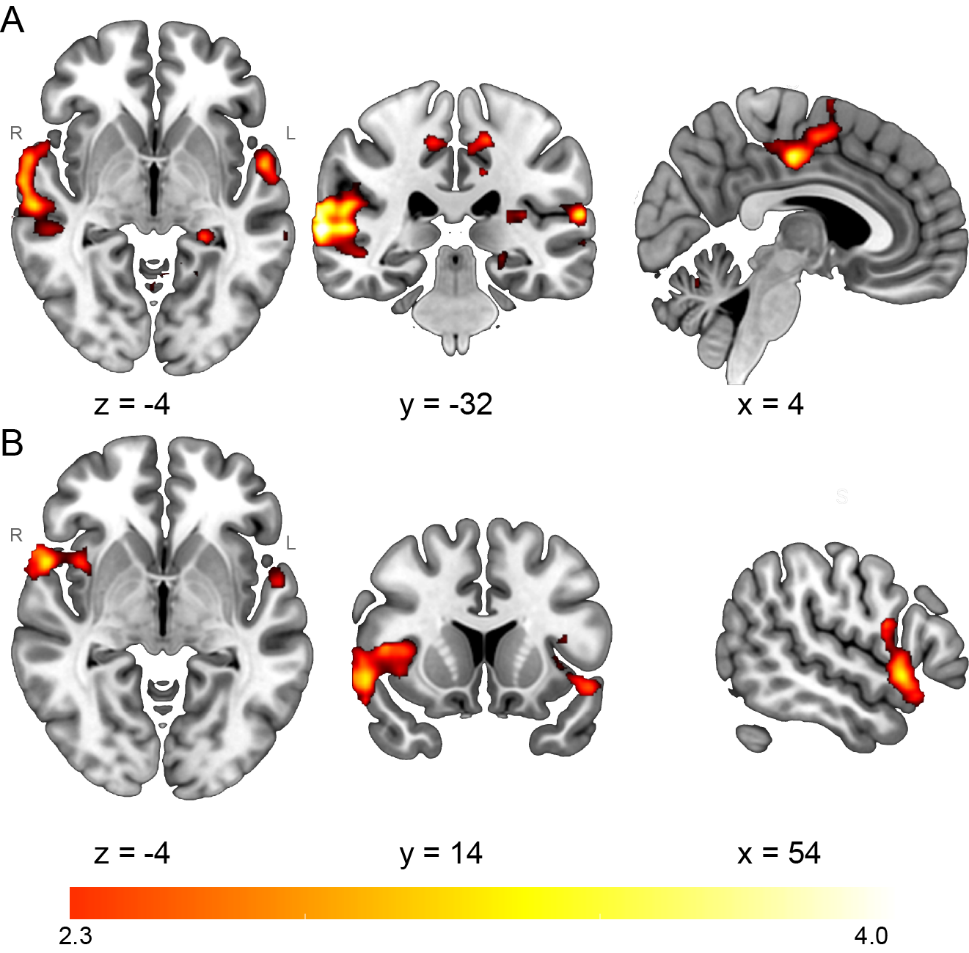

### References

1.	First M, Spitzer, R., Gibbon, M. & Williams, J. Structured Clinical Interview for DSM-IV Axis I Disorders. Biometrics Research; 2002.

2.	Nelis S, Holmes EA, Griffith JW, Raes F. Mental imagery during daily life: Psychometric evaluation of the Spontaneous Use of Imagery Scale (SUIS). *Psychol Belg*. Jan 20 2014;54(1):19-32. doi:10.5334/pb.ag

3.	Marks DF. Visual imagery differences in the recall of pictures. *Br J Psychol*. Feb 1973;64(1):17-24. doi:10.1111/j.2044-8295.1973.tb01322.x

4.	Andrade J, May J, Deeprose C, Baugh SJ, Ganis G. Assessing vividness of mental imagery: The Plymouth Sensory Imagery Questionnaire. *Br J Psychol*. Nov 2014;105(4):547-63. doi:10.1111/bjop.12050

5.	O'Donnell C, Di Simplicio M, Burnett Heyes S. Hypomanic-like experiences and spontaneous emotional mental imagery. *J Affect Disord*. Dec 1 2020;277:742-746. doi:10.1016/j.jad.2020.08.003

6.	Deeprose C, Holmes EA. An exploration of prospective imagery: the impact of future events scale. *Behav Cogn Psychother*. Mar 2010;38(2):201-9. doi:10.1017/S1352465809990671

7.	Altman EG, Hedeker D, Peterson JL, Davis JM. The Altman Self-Rating Mania Scale. *Biol Psychiatry*. Nov 15 1997;42(10):948-55. doi:10.1016/S0006-3223(96)00548-3

8.	Rush AJ, Trivedi MH, Ibrahim HM, et al. The 16-Item Quick Inventory of Depressive Symptomatology (QIDS), clinician rating (QIDS-C), and self-report (QIDS-SR): a psychometric evaluation in patients with chronic major depression. *Biol Psychiatry*. Sep 1 2003;54(5):573-83. doi:10.1016/s0006-3223(02)01866-8

9.	Beck AT, Epstein N, Brown G, Steer RA. An inventory for measuring clinical anxiety: psychometric properties. *J Consult Clin Psychol*. Dec 1988;56(6):893-7. doi:10.1037//0022-006x.56.6.893

10.	Henry C, M'Bailara K, Lepine JP, Lajnef M, Leboyer M. Defining bipolar mood states with quantitative measurement of inhibition/activation and emotional reactivity. *J Affect Disord*. Dec 2010;127(1-3):300-4. doi:10.1016/j.jad.2010.04.028

11.	Di Simplicio M, Holmes EA, Rathbone CJ. Self-images in the present and future: Role of affect and the bipolar phenotype. *J Affect Disord*. Nov 15 2015;187:97-100. doi:10.1016/j.jad.2015.08.042

12.	Jenkinson M, Bannister P, Brady M, Smith S. Improved optimization for the robust and accurate linear registration and motion correction of brain images. *Neuroimage*. Oct 2002;17(2):825-41. doi:10.1016/s1053-8119(02)91132-8

13.	Smith SM. Fast robust automated brain extraction. *Hum Brain Mapp*. Nov 2002;17(3):143-55. doi:10.1002/hbm.10062

14.	Beckmann CF, Smith SM. Probabilistic independent component analysis for functional magnetic resonance imaging. *IEEE Trans Med Imaging*. Feb 2004;23(2):137-52. doi:10.1109/TMI.2003.822821

15.	Griffanti L, Douaud G, Bijsterbosch J, et al. Hand classification of fMRI ICA noise components. *Neuroimage*. Jul 1 2017;154:188-205. doi:10.1016/j.neuroimage.2016.12.036

16.	Greve DN, Fischl B. Accurate and robust brain image alignment using boundary-based registration. *Neuroimage*. Oct 15 2009;48(1):63-72. doi:10.1016/j.neuroimage.2009.06.060

17.	Jenkinson M, Smith S. A global optimisation method for robust affine registration of brain images. *Med Image Anal*. Jun 2001;5(2):143-56. doi:10.1016/s1361-8415(01)00036-6

18.	Jenkinson M, Beckmann CF, Behrens TE, Woolrich MW, Smith SM. Fsl. *Neuroimage*. Aug 15 2012;62(2):782-90. doi:10.1016/j.neuroimage.2011.09.015

19.	Salimi-Khorshidi G, Douaud G, Beckmann CF, Glasser MF, Griffanti L, Smith SM. Automatic denoising of functional MRI data: combining independent component analysis and hierarchical fusion of classifiers. *Neuroimage*. Apr 15 2014;90:449-68. doi:10.1016/j.neuroimage.2013.11.046

20.	Woolrich MW, Ripley BD, Brady M, Smith SM. Temporal autocorrelation in univariate linear modeling of FMRI data. *Neuroimage*. Dec 2001;14(6):1370-86. doi:10.1006/nimg.2001.0931

21.	Filippini N, MacIntosh BJ, Hough MG, et al. Distinct patterns of brain activity in young carriers of the APOE-epsilon4 allele. *Proc Natl Acad Sci U S A*. Apr 28 2009;106(17):7209-14. doi:10.1073/pnas.0811879106

22.	Winkler AM, Ridgway GR, Webster MA, Smith SM, Nichols TE. Permutation inference for the general linear model. *Neuroimage*. May 15 2014;92:381-97. doi:10.1016/j.neuroimage.2014.01.060
